# Optical coherence tomography assessment of axonal and neuronal damage of the retina in patients with familial and sporadic multiple sclerosis

**DOI:** 10.1101/2022.04.20.22274092

**Authors:** M Grudziecka Pyrek, K Selmaj

## Abstract

**Objective:** Optical coherence tomography (OCT) assessment of axonal and neuronal damage of the retina in patients with familial and sporadic multiple sclerosis.

**Methods:** 45 patients with familial MS (fMS), 58 patients with sporadic MS (sMS) and 35 healthy controls were included in the study. OCT was performed with the spectral domain optical coherence tomography (SD-OCT, Heidelberg Engineering, Germany). The retinal nerve fiber layer (RNFL) thickness and macular volume (MV) were measured.

**Results:** A significant thinning of the global RNFL thickness was detected in both forms of MS compared to control group, (86,61 (+/- 14,74) µm in sMS, 85,8 (+/- 12,7) µm in fMS, 97,96 (+/- 7,6) µm in control group; p <0,001). There was no significant difference in the global RNFL thickness between sMS and fMS. A significant reduction of the MV was shown in sMS and fMS compared to control group (8,12 (+/- 1,14) mm^3^ in sMS, 8,1 (+/- 1,12) mm^3^ in fMS, and 8,81 (+/- 0,31) mm^3^ in control group; p = 0,003). No difference in MV between sMS and fMS was found. However, in eyes with history of optic neuritis (ON) MV was significantly reduced in sMS versus fMS (8,12 (+/- 2,87) mm^3^ vs. 8,42 (+/- 0,54) mm^3^; p=0,05).

**Conclusion:** We confirmed the presence of axonal and neuronal damage of the retina in sMS and fMS. ON induced a significantly greater reduction of MV in sMS compared to fMS, indicating a stronger neuronal damage in ON eyes in sMS than in fMS.

**KEY MESSAGES:** The familial MS accounts for a significant proportion of MS patients and there is still ongoing discussion on the distinction between familial (fMS) and sporadic (sMS) forms of this disease. Using OCT, we confirmed the presence of axonal and neuronal damage in the retina in both forms of MS. We found that optic neuritis (ON) induced a greater retinal neuronal damage in sMS than in fMS. These results support the conclusion that there are some discrete differences in pathological processes occurring in the retina in sMS and fMS.

## INTRODUCTION

Multiple sclerosis (MS) is the most common chronic inflammatory demyelinating disorder of the central nervous system (CNS). Two major forms of MS have been recognized: sporadic (sMS) and familial (fMS). In fMS apart from an index case at least one more family member should be diagnosed with MS,[1]. It is estimated that fMS affects about 12.6 - 20% of all patients with MS (pwMS),[2-4]. The genetics of MS is complex and correspond to polygenic trait. It is believed that MS genetics can explain up to half of the disease’s heritability,[5, 6]. Classical twin studies showed that unaffected monozygotic twin have much higher risk to develop MS, 25-30%, than dizygotic twin, 2-5%,[7, 8]. The average risk of developing MS by relatives of pwMS ranged from 3 to 5%, with the highest risk for first-degree relatives, and was 30 to 50 times higher than the 0,1% risk for the general population,[2]. These data suggest that fMS might represent a more genetically driven form of MS versus sMS. Clinical phenotyping of these two forms of MS did not reveal any major differences,[9-11]. However, some discordance was observed. It was found that pwMS with multiple affected relatives had a higher incidence of optic neuritis (ON) as the first relapse, a lower risk of another relapse in the first year of disease, a longer interval between the first and second relapse, and a longer time to permanent neurological deficit,[3]. Also MRI studies have shown some differences between these two forms of MS. In fMS compared to sMS was observed a larger T1-lesion volume and a trend toward lower MTR of T1-lesions,[12]. Our own earlier studies showed that MTR abnormalities were more widespread in fMS than in sMS. MTR values were reduced mainly in the corpus callosum and in the cerebral and cerebellar peduncles, primarily involving areas of highly myelinated white matter,[13]. Proton magnetic resonance spectroscopy showed a slight decrease in NAA/Cho and NAA/Cr ratios in normal appearing white matter in sMS versus fMS, whereas Cho/Cr ratio showed an increased trend. These results might indicate more pronounced neurodegenerative injury in sMS than in fMS,[14].

To address this hypothesis, we applied optic coherence tomography (OCT) technique and aimed to study axonal and neuronal damage of the retina in patients with sMS and fMS. OCT is a non-invasive interferometric technique, allowing for assessment of axonal and neuronal damage by measurement of retinal nerve fibre layer (RNFL) thickness and macular volume (MV). The RNFL is formed by unmyelinated retinal ganglion cell axons, whereas ganglion cell neurons are a major component of the macula,[15, 16]. Several studies reported a positive correlation of the RNFL thinning with brain atrophy in MS, which suggested that RNFL thickness might be an indirect marker of diffuse axonal damage in the brain,[17-19]. The reduction of MV in MS was associated with neurodegenerative neuronal changes. Hence, MV might represent a quantitative marker of neuronal damage in the brain,[20-22].

We analysed RNFL thickness and MV in a cohort of patients with fMS and sMS and healthy volunteers and investigated how ON affected the OCT parameters in patients with both forms of MS.

## METHODS

### Study design

In this prospective study, we analysed data from patients with relapsing-remitting multiple sclerosis (pwRRMS) and healthy volunteers of University Hospital in Lodz, Poland, between 2012 and 2016. All study participants had OCT examination with RNFL and MV measurements of both eyes. Two groups of pwRRMS patients were analysed, sporadic MS patients (sMS) and patients with familial history of MS (fMS). To evaluate the effect of ON on OCT parameters in sMS and fMS, the examined eyes were divided into three subgroups: eyes with a history of ON, eyes contralateral to eyes with history of ON (Cont-L) and eyes of patients without history of ON (Non-ON).

The OCT measurements were compared between both MS forms and with healthy volunteers.

### Participants

A total of 103 pwRRMS and 33 healthy volunteers, aged 18 – 60, were included in the study. 58 patients had sMS and 45 fMS. MS diagnosis was confirmed according to the McDonald 2010 criteria,[23]. Patients with fMS required to have at least one more MS case within relatives of kinship degree 1-3.

We excluded patients with any other neurological disorder, patients with optic neuritis (ON) 6 months prior to study entry and patients with other ophthalmic disorders that might affect OCT measurements, such as glaucoma, optic nerve damage, retinopathy, severe myopia (greater than 6.00 dioptres). The other exclusion criteria involved medication with intraocular steroids, intraocular anti-angiogenic drugs and chronic systemic steroids.

The study was approved by Ethical Commission of the Medical University of Lodz; Ethic Approval/Registration Numbers: RNN/83/13/KE, RNN/178/16/KE. All study participants gave written informed consent to participate in the study.

### Optical coherence tomography (OCT)

OCT was performed by a certified operator on a spectral domain OCT device in a darkened room without using of pharmacological pupil dilators (Heidelberg Spectralis OCT software version 4.0.0.0.0). A circular RNFL thickness scan was measured, centred on the optic nerve disc. The result was shown as a circular diagram with automatically calculated global RNFL thickness and RNFL thickness in 4 quadrants: nasal, temporal, superior and inferior and in the papillomacular bundle (PMB), representing the middle 30 degrees of the temporal quadrant, relative to a normative database of age, race, and sex. The macula area was scanned in 25 sections in 240 µm inter-scan distance. A 6-mm diameter circular macular volume scan was performed with the centre located in the fovea. OCT software automatically calculated total macular volume of the entire scanned area.

### Expanded Disability Status Scale (EDSS)

Study participants were neurologically examined and their disability was assessed according to the Expanded Disability Status Scale (EDSS), which includes 8 functional systems: pyramidal, cerebellar, brainstem, sensory, bowel and bladder, visual, cerebral and other neurologic findings attributed to MS,[24]. In each functional system, the patient received a certain point value from which the final EDSS score was then calculated.

### Statistical analysis

Shapiro-Wilk and D’Agostino-Pearson tests were used to assess the normality of distributions. Student’s t test and Mann-Whitney test (after Fisher-Snedecor test) were used to compare OCT parameters between both groups of pwRRMS and healthy controls. The analysis of variance (ANOVA) method was used to compare multiple groups. A p value of <0.05 was considered significant. We performed all statistical analyses using the MedCalc software (V.18.2).

## RESULTS

### Patients characteristics

Demographic and clinical data are shown in Table 1. The median disease duration in sMS and fMS was 8,27 (+/-7,56) and 11,2 (+/-8,2) years. In sMS, 36 patients were females and 22 were men, and in fMS the gender distribution was 34 and 11, respectively. The mean EDSS was 1,68 (+/-1,23) in sMS and 2,1 (+/-1,07) in fMS. The number of relapses in the last 2 years before OCT examination in sMS was 1,1 (+/-0,83) and in fMS 1,07 (+/-0,96). The percentage of patients with previous ON was 43% in sMS and 56% in fMS. The mean time from ON was 8,55 (+/-7,69) years in sMS and 10,24 (+/-7,56) year.

**Table 1.** Demographics of study participants

|  | fMS | sMS | Control group |
| --- | --- | --- | --- |
| Number of participants in the study | 45 | 58 | 33 |
| Age (+/- SD) | 39,82 (9,5) | 38,28(10,49) | 36,76 (9,11) |
| Number of women | 34 | 36 | 23 |
| Number of men | 11 | 22 | 10 |
| Duration of MS (years) (+/- SD) | 11,2 (8,2) | 8,27 (7,56) |  |
| EDSS (+/- SD) | 2,1 (1,07) | 1,68 (1,23) |  |
| Number of relapses in the last 2 years before OCT examination | 1,07 (0,96) | 1,1 (0,83) |  |
| Percentage of patients with a history of ON | 56% | 43% |  |
| Time since ON (years), (+/- SD) | 10,24 (6,96) | 8,55 (7,69) |  |

fMS, familial multiple sclerosis; sMS, sporadic multiple sclerosis; ON, optic neuritis; SD, standard deviation

### Comparison of RNFL thickness between sMS, fMS and control group

The global RNFL thickness in sMS was 86,61 (+/- 14,74) μm, and in fMS 85,8 (+/- 12,7) μm. For both sMS and fMS, these values were significantly lower compared to the control group, in which the global RNFL thickness was 97,96 (+/- 7,6) μm, (p<0,001 for both forms of pwRRMS). Segmental analysis showed the RNFL thinning in all segments in sMS and fMS compared to control group, reaching the greatest differences in the papillomacular bundle (PMB), temporal and inferior segments (p<0,001). Comparative analysis of sMS with fMS showed no significant difference in global and segmental RNFL thickness (Table 2).

**Table 2.** Comparison of RNFL thickness (µm) between sMS, fMS and control group

|  | OCT parameters (+/-SD) | p - value |
| --- | --- | --- |

|  | fMS | sMS | control group | fMS vs. sMS | fMS vs. control group | sMS vs. control group |
| --- | --- | --- | --- | --- | --- | --- |
| Global RNFL | 85,8<br>(12,7) | 86,61<br>(14,74) | 97,95<br>(7,6) | 0,77 | <0,001 | <0,001 |
| RNFL in the temporal segment | 54,98<br>(1,12) | 56,95<br>(14,98) | 68,32<br>(9,43) | 0,48 | <0,001 | <0,001 |
| RNFL in the papillomacular bundle (PMB) | 41,19<br>(10,02) | 43,98<br>(11,1) | 52,67<br>(7,53) | 0,19 | <0,001 | <0,001 |
| RNFL in the superior segment | 222,26<br>(32,15) | 222,06<br>(40,24) | 245,35<br>(21,37) | 0,98 | 0,004 | 0,004 |
| RNFL in the nasal segment | 66,21<br>(13,56) | 65,94<br>(13,43) | 72,12<br>(10,56) | 0,92 | 0,062 | 0,062 |
| RNFL in the inferior segment | 230,48<br>(32,44) | 225,01<br>(40,71) | 257,67<br>(24,91) | 0,46 | <0,001 | <0,001 |

RNFL, retinal nerve fiber layer; fMS, familial multiple sclerosis; sMS, sporadic multiple sclerosis; SD, standard deviation

### Comparison of MV between sMS, fMS and control group

The MV was significantly lower in sMS and fMS patients compared with controls (8,12 (+/- 1,14) mm^3^ and 8,1 (+/- 1,12) mm^3^ vs. 8,81 (+/- 0,31) mm^3^, respectively), p=0,003. There was no significant difference in the MV between sMS and fMS (Table 3).

**Table 3.** Comparison of macular volume (mm^3^) between sMS, fMS and control group

|  | OCT parameters (+/-SD) |  |  | p-value |  |  |
| --- | --- | --- | --- | --- | --- | --- |
|  | fMS | sMS | control group | fMS vs. sMS | fMS vs. control group | sMS vs. control group |
| MV | 8,1<br>(1,12) | 8,12<br>(1,14) | 8,81<br>(0,31) | 0,96 | 0,003 | 0,003 |

MV, macular volume; fMS, familial multiple sclerosis; sMS, sporadic multiple sclerosis; SD, standard deviation

### Impact of ON on RFNL in sMS and fMS

RNFL was measured in eyes with history of optic neuritis (ON) in sMS (n=33) and in fMS (n=36), in eyes contralateral to eyes with history of ON (Cont-L) in sMS (n=17) and in fMS (n=14), in eyes of patients without history of optic neuritis (Non-ON), in sMS (n= 66) in and in fMS (n=40), and in eyes from control subjects (n= 66). For both sMS and fMS the RNFL thickness in ON vs. Non-ON eyes was significantly lower in ON. Similarly RNFL in ON vs. control eyes in sMS and fMS was significantly lower in ON (Table 4). When directly comparing, how ON affected RNFL in sMS and fMS we found no significant difference in RFNL thickness of ON eyes between sMS and fMS, p=0,37. The only observed trend for thinner RNFL occurred in papillomacular bundle in patients with fMS, p=0,09 (Table 5).

**Table 4.** Impact of ON on global RNFL thickness (µm) in sMS and fMS

|  | **OCT parameters in sMS**<br>(+/-SD) |  |  |  | **p - value** |  |  |  |  |  |
| --- | --- | --- | --- | --- | --- | --- | --- | --- | --- | --- |
| Eyes | Non-ON | Cont-L | ON | control group | Non-ON vs. Cont-L | Non-ON vs. ON | Cont-L vs. ON | Non-ON vs. control group | Cont-L vs. control group | ON vs. control group |
| Global RNFL | 91,46<br>(13,36) | 90,29<br>(12,39) | 78<br>(10,86) | 97,95<br>(7,6) | 0,24 | <0,001 | <0,001 | 0,02 | 0,0007 | <0,0001 |
|  | **OCT parameters in fMS**<br>(+/-SD) |  |  |  | **p - value** |  |  |  |  |  |
| Eyes | Non-ON | Cont-L | ON | control group | Non-ON vs. Cont-L | Non-ON vs. ON | Cont-L vs. ON | Non-ON vs. control group | Cont-L vs. control group | ON vs. control group |
| Global RNFL | 91,28<br>(9,37) | 89,29<br>(13,03) | 80,86<br>(13,58) | 97,95<br>(7,6) | 0,53 | <0,001 | 0,21 | 0,056 | 0,001 | <0,0001 |

RNFL, retinal nerve fiber layer; fMS, familial multiple sclerosis; sMS, sporadic multiple sclerosis; SD, standard deviation; Non-ON, eye of patient without history of optic neuritis; Cont-L, eye contralateral to eye with history of ON; ON, eye with history of ON

**Table 5.** Comparison of RNFL thickness between fMS and sMS in different subgroups of eyes

| OCT parameters | p-value<br>fMS vs. sMS |  |  |
| --- | --- | --- | --- |
|  | Non-ON | Cont-L | ON |
| Global RNFL | 0,94 | 0,83 | 0,37 |
| RNFL in the temporal segment | 0,57 | 0,27 | 0,53 |
| RNFL in the papillomacular bundle (PMB) | 0,83 | 0,31 | 0,09 |
| RNFL in the superior segment | 0,29 | 0,89 | 0,17 |
| RNFL in the nasal segment | 0,91 | 0,8 | 0,5 |
| RNFL in the inferior segment | 0,49 | 0,67 | 0,22 |

RNFL, retinal nerve fiber layer; fMS, familial multiple sclerosis; sMS, sporadic multiple sclerosis; Non-ON, eye of patient without history of optic neuritis; Cont-L, eye contralateral to eye with history of ON; ON, eye with history of ON

### Impact of ON on MV in sMS and fMS

MV was measured in eyes with ON in sMS (n=33) and in fMS (n=36), in Cont-L eyes in sMS (n=17) and in fMS (n=14), in Non-ON eyes in sMS (n= 66) and in fMS (n=40) and in eyes from control subjects (n= 66). For sMS the MV was significantly lower in ON than in Non-ON and control eyes, p<0,001 and p=0,0003, respectively. For fMS, MV in ON eyes was significantly lower only in comparison to control eyes, p=0,007 (Table 6), and did not differ from Non-ON eyes. Most importantly, when directly comparing MV in sMS with fMS it was found that MV in sMS was significantly reduced versus fMS, p=0,05 (Table 7).

**Table 6.** Impact of ON on MV (mm^3^) in sMS and fMS

|  | **OCT parameters in sMS**<br>**(+/-SD)** |  |  |  | **p-value** |  |  |  |  |  |
| --- | --- | --- | --- | --- | --- | --- | --- | --- | --- | --- |
| Eyes | Non-ON | Cont-L | ON | control group | Non-ON vs. Cont-L | Non-ON vs. ON | Cont-L vs. ON | Non-ON vs. control group | Cont-L vs. control group | ON vs. control group |
| MV | 8,59<br>(1,55) | 8,45<br>(2,83) | 8,12<br>(2,87) | 8,81<br>(0,31) | 0,37 | <0,001 | 0,07 | 0,02 | 0,008 | 0,0003 |
|  | **OCT parameters in fMS**<br>**(+/-SD)** |  |  |  | **p-value** |  |  |  |  |  |
| Eyes | Non-ON | Cont-L | ON | control group | Non-ON vs. Cont-L | Non-ON vs. ON | Cont-L vs. ON | Non-ON vs. control group | Cont-L vs. control group | ON vs. control group |
| MV | 8,46<br>(0,42) | 8,48<br>(0,34) | 8,42<br>(0,54) | 8,81<br>(0,31) | 0,67 | 0,83 | 0,74 | 0,002 | 0,04 | 0,007 |

MV, macular volume; fMS, familial multiple sclerosis; sMS, sporadic multiple sclerosis; Non-ON, eye of patient without history of optic neuritis; Cont-L, eye contralateral to eye with history of ON; ON, eye with history of ON

**Table 7.** Comparison of MV (mm^3^) between fMS and sMS in different subgroups of eyes

| **OCT parameters** | **p-value**<br>fMS vs. sMS |  |  |
| --- | --- | --- | --- |
|  | Non-ON | Cont-L | ON |
| MV | 0,12 | 0,88 | 0,05 |

MV, macular volume; fMS, familial multiple sclerosis; sMS, sporadic multiple sclerosis; Non-ON, eye of patient without history of optic neuritis; Cont-L, eye contralateral to eye with history of ON; ON, eye with history of ON

## DISCUSSION

We have found that RNFL thickness and MV in sMS and fMS were significantly diminished in comparison to control subjects. These findings confirmed the presence of axonal and neuronal damage of the retina in patients with sMS and fMS. The global and segmental thickness of RFNL and MV were similar in both sMS and fMS. However, we have found that the impact of previous optic neuritis on MV was bigger in sMS than in fMS, suggesting more pronounced neuronal damage of the retina in sMS.

OCT offers non-invasive and relatively simple method to assess axonal and neuronal changes of the retina. RNFL measures axonal thickness of ganglion cells and MV corresponds to the count of two or more layers of ganglion cells,[16, 17, 25]. Over the last several years numerous studies demonstrated diminished RNFL in pwMS,[26-28]. Most importantly, RFNL thickness correlated to diffuse axonal changes throughout the CNS of pwMS,[18,19]. In particular RNFL correlated with global and regional brain atrophy,[18-22], with MRI lesion load,[18, 29], MTR measures,[30], with NAA/Cho ratio in proton brain MRI spectroscopy,[31], as well as with cerebrospinal fluid neurofilament light chain levels,[32]. On the other hand measurements of MV and ganglion cell-inner plexiform layer (GCIPL) thickness were suggested to be more reliable at detecting neuronal degenerative processes in MS,[26, 28]. Similarly to RNFL thinning in pwMS reduction of MV and GCIPL thickness correlated with brain pathology and enhanced brain atrophy,[20,22].

The unique ability to monitor axonal and neuronal injury *in vivo* using OCT contributed to better understanding of global pathological processes in MS. The reported correlation of RNFL thickness and MV with MRI parameters, particularly with markers of brain atrophy in pwMS, makes OCT a useful method for monitoring neurodegenerative changes in this disease,[19, 22]. The findings that ON induced a larger reduction of MV in patients with sMS should be interpreted from the mechanistic point of view how ON affects the retinal pathology. Changes in the retina caused by inflammation in the optic nerve in the classical interpretation represents a consequence of its demyelination, leading to retrograde axonopathy as thinning of RNFL. This pattern of RNFL changes we observed in both sMS and fMS. However, in sMS ON induced significantly greater reduction of MV than in fMS and in this latter form reduction of MV was quite limited. Since MV corresponds to number of neuronal ganglion cells and it is recognized as a marker for neuronal cell pathology,[20-22, 34] one might suggest that ON in sMS has a particular impact on neuronal damage of the retina. This observation might be of interest from the perspective of the role of primary neuronal damage in the CNS of pwMS. It has been suggested that MS pathology apart from demyelination and secondary axonopathy also involves neuronal damage,[35, 36]. Its relationship to inflammation is still under discussion. Differences in the degree of neuronal damage in the retina after ON between sMS and fMS might suggest discordance in inflammation-induced tissue damage in these two forms of MS. Whether this would be related to specific genetic background in sMS and fMS remains for further investigations. Nevertheless these results might indicate that retinal changes in pwMS might have more complex pattern influenced by genetic trait.

There is still ongoing discussion on the distinction between familial and sporadic forms of MS. The major issue is how much these two forms might differ in terms of genetic background leading to some differences in pathologic mechanisms. The attempt to characterize fMS by genetic screening is not available yet. However, in that context, of interest might be results of studies on MRI findings in asymptomatic relatives of patients with sMS and fMS. The 11% of asymptomatic siblings of MS patients showed demyelinating brain lesions similar to that seen in MS,[37]. Our own earlier study with MTR assessment of normal appearing white matter reported diminished MTR histogram peak heights in asymptomatic relatives of MS patients,[38]. Similarly a study with proton magnetic resonance spectroscopy showed lower NAA/Cho and higher Cho/Cr ratio in asymptomatic relatives of pwMS,[15]. In the large cohort of first degree asymptomatic relatives of sMS and fMS patients a higher prevalence of MRI lesions was found in fMS, 10% compared with 4% in sMS,[39]. The demonstrated differences in brain imaging in relatives of patients with fMS and sMS may perhaps support the existence of some genetic differences between these two forms of MS.

Our results have some obvious limitations related to the definition of fMS. However, the cohort of pwMS used in this study was derived from prospective analysis reducing biases related to the development of new cases in relatives over time. In addition the mean age of pwMS in this study, 39,86 for fMS and 38,28 years for sMS, was clearly above pooled mean age of fMS onset of 15 studies reported as 28,7Lyears,[40]. Also, the affected members of the family were carefully evaluated with medical records and neurological examinations.

In conclusion, our findings on a different impact of ON on MV in sMS and fMS and more pronounced neuronal injury of the retina in sMS patients might support discrete differences in pathologic mechanisms between sMS and fMS.

## Data Availability

All data produced in the present work are contained in the manuscript

## Contributors

MGP contributed to data collection, analysed imaging, designed and conceptualised the study, analysed and interpreted all data, and drafted the manuscript. KS designed and conceptualised the study, analysed and interpreted all data, drafted, revised the manuscript. KS was the guarantor for this study.

The corresponding author attests that all listed authors meet authorship criteria and that no others meeting the criteria have been omitted

## Copyright/licence for publication

*The Corresponding Author has the right to grant on behalf of all authors and does grant on behalf of all authors, to the Publishers and its licensees in perpetuity, in all forms, formats and media (whether known now or created in the future), to i) publish, reproduce, distribute, display and store the Contribution, ii) translate the Contribution into other languages, create adaptations, reprints, include within collections and create summaries, extracts and/or, abstracts of the Contribution, iii) create any other derivative work(s) based on the Contribution, iv) to exploit all subsidiary rights in the Contribution, v) the inclusion of electronic links from the Contribution to third party material where-ever it may be located; and, vi) licence any third party to do any or all of the above*.

## Patient consent for publication

Not applicable.

## Ethics approval

The study was approved by Ethical Commission of the Medical University of Lodz, Poland (registration numbers: RNN/83/13/KE, RNN/178/16/KE).

## Provenance and peer review

Not commissioned; externally peer reviewed.

## Data availability statement

Data are available upon reasonable request.

## REFERENCES

1. Hartung HP, Will RG, Francis D, et al. Familial multiple sclerosis. J Neurol Sci. 1988;83(2-3):259–268. doi:10.1016/0022-510x(88)90073-1

2. Sadovnick AD, Baird PA. The familial nature of multiple sclerosis: age-corrected empiric recurrence risks for children and siblings of patients. Neurology. 1988;38(6):990–991. doi:10.1212/wnl.38.6.990

3. Ebers GC, Koopman WJ, Hader W, et al. The natural history of multiple sclerosis: a geographically based study: 8: familial multiple sclerosis. Brain. 2000;123 Pt 3:641–649. doi:10.1093/brain/123.3.641

4. Harirchian MH, Fatehi F, Sarraf P, Honarvar NM, Bitarafan S. Worldwide prevalence of familial multiple sclerosis: A systematic review and meta-analysis. Mult Scler Relat Disord. 2018;20:43–47. doi:10.1016/j.msard.2017.12.015

5. Sawcer S. The complex genetics of multiple sclerosis: pitfalls and prospects. Brain. 2008;131(Pt 12):3118–3131. doi:10.1093/brain/awn081

6. Patsopoulos NA, De Jager PL. Genetic and gene expression signatures in multiple sclerosis. Mult Scler. 2020;26(5):576–581. doi:10.1177/1352458519898332

7. Sadovnick AD, Armstrong H, Rice GP, et al. A population-based study of multiple sclerosis in twins: update. Ann Neurol. 1993;33(3):281–285. doi:10.1002/ana.410330309

8. Hansen T, Skytthe A, Stenager E, Petersen HC, Brønnum-Hansen H, Kyvik KO. Concordance for multiple sclerosis in Danish twins: an update of a nationwide study. Mult Scler. 2005;11(5):504–510. doi:10.1191/1352458505ms1220oa

9. Dyment DA, Cader MZ, Willer CJ, Risch N, Sadovnick AD, Ebers GC. A multigenerational family with multiple sclerosis. Brain. 2002;125(Pt 7):1474–1482. doi:10.1093/brain/awf158

10. Weinshenker BG, Bulman D, Carriere W, Baskerville J, Ebers GC. A comparison of sporadic and familial multiple sclerosis. Neurology. 1990;40(9):1354–1358. doi:10.1212/wnl.40.9.1354

11. Ceccarelli A, Mifsud VA, Dogar A. Demographic and clinical characteristics of familial and sporadic multiple sclerosis: A single center exploratory study from Abu Dhabi. J Clin Neurosci. 2020;76:145–147. doi:10.1016/j.jocn.2020.04.007

12. Tipirneni A, Weinstock-Guttman B, Ramanathan M, et al. MRI characteristics of familial and sporadic multiple sclerosis patients. Mult Scler. 2013;19(9):1145–1152. doi:10.1177/1352458512469697

13. Siger-Zajdel M, Selmaj K. Magnetisation transfer ratio analysis of normal appearing white matter in patients with familial and sporadic multiple sclerosis. J Neurol Neurosurg Psychiatry. 2001;71(6):752–756. doi:10.1136/jnnp.71.6.752

14. Siger-Zajdel M, Selmaj K. Proton magnetic resonance spectroscopy of normal appearing white matter in asymptomatic relatives of multiple sclerosis patients. Eur J Neurol. 2006;13(3):296–298. doi:10.1111/j.1468-1331.2006.01177.x

15. Yanoff M, Duker J, Augsburger J. Ophthalmology. Mosby, 2003

16. Duker J, Waheed N, Goldman D. Handbook of Retinal OCT: Optical Coherence Tomography. Elsevier, 2014

17. Trip SA, Schlottmann PG, Jones SJ, et al. Optic nerve atrophy and retinal nerve fibre layer thinning following optic neuritis: evidence that axonal loss is a substrate of MRI-detected atrophy. Neuroimage.2006;31(1):286–293. doi:10.1016/j.neuroimage.2005.11.051

18. Siger M, Dziegielewski K, Jasek L, et al. Optical coherence tomography in multiple sclerosis: thickness of the retinal nerve fiber layer as a potential measure of axonal loss and brain atrophy. J Neurol. 2008;255(10):1555–1560. doi:10.1007/s00415-008-0985-5

19. Abalo-Lojo JM, Limeres CC, Gómez MA, et al. Retinal nerve fiber layer thickness, brain atrophy, and disability in multiple sclerosis patients. J Neuroophthalmol. 2014;34(1):23–28. doi:10.1097/WNO.0000000000000057

20. Dörr J, Wernecke KD, Bock M, et al. Association of retinal and macular damage with brain atrophy in multiple sclerosis. PLoS One. 2011;6(4):e18132. Published 2011 Apr 8. doi:10.1371/journal.pone.0018132

21. Burkholder BM, Osborne B, Loguidice MJ, et al. Macular volume determined by optical coherence tomography as a measure of neuronal loss in multiple sclerosis. Arch Neurol. 2009;66(11):1366–1372. doi:10.1001/archneurol.2009.230

22. Saidha S, Al-Louzi O, Ratchford JN, et al. Optical coherence tomography reflects brain atrophy in multiple sclerosis: A four-year study. Ann Neurol. 2015;78(5):801–813. doi:10.1002/ana.24487

23. Polman CH, Reingold SC, Banwell B, et al. Diagnostic criteria for multiple sclerosis: 2010 revisions to the McDonald criteria. Ann Neurol. 2011;69(2):292–302. doi:10.1002/ana.22366

24. Meyer-Moock S, Feng YS, Maeurer M, Dippel FW, Kohlmann T. Systematic literature review and validity evaluation of the Expanded Disability Status Scale (EDSS) and the Multiple Sclerosis Functional Composite (MSFC) in patients with multiple sclerosis. BMC Neurol. 2014;14:58. Published 2014 Mar 25. doi:10.1186/1471-2377-14-58

25. Staurenghi G, Sadda S, Chakravarthy U, Spaide RF; International Nomenclature for Optical Coherence Tomography (IN•OCT) Panel. Proposed lexicon for anatomic landmarks in normal posterior segment spectral-domain optical coherence tomography: the IN•OCT consensus. Ophthalmology. 2014;121(8):1572–1578. doi:10.1016/j.ophtha.2014.02.023

26. Talman LS, Bisker ER, Sackel DJ, et al. Longitudinal study of vision and retinal nerve fiber layer thickness in multiple sclerosis. Ann Neurol. 2010;67(6):749–760. doi:10.1002/ana.22005

27. Costello F, Coupland S, Hodge W, et al. Quantifying axonal loss after optic neuritis with optical coherence tomography. Ann Neurol. 2006;59(6):963–969. doi:10.1002/ana.20851

28. Petzold A, Balcer LJ, Calabresi PA, et al. Retinal layer segmentation in multiple sclerosis: a systematic review and meta-analysis. Lancet Neurol. 2017;16(10):797–812. doi:10.1016/S1474-4422(17)30278-8

29. Cilingir V, Batur M, Bulut MD, Milanlioglu A, Yılgor A, Batur A, Yasar T, Tombul T. The association between retinal nerve fibre layer thickness and corpus callosum index in different clinical subtypes of multiple sclerosis. Neurol Sci. 2017 Jul;38(7):1223–1232.

30. Trip SA, Schlottmann PG, Jones SJ, et al. Optic nerve magnetization transfer imaging and measures of axonal loss and demyelination in optic neuritis. Mult Scler. 2007;13(7):875–879. doi:10.1177/1352458507076952

31. Pfueller CF, Brandt AU, Schubert F, et al. Metabolic changes in the visual cortex are linked to retinal nerve fiber layer thinning in multiple sclerosis. PLoS One. 2011;6(4):e18019. Published 2011 Apr 6. doi:10.1371/journal.pone.0018019

32. Modvig S, Degn M, Sander B et al. Cerebrospinal fluid neurofilament light chain levels predict visual outcome after optic neuritis. Mult Scler. 2016;22(5):590–8. doi: 10.1177/1352458515599074

33. Petzold A, Balcer LJ, Calabresi PA, et al. Retinal layer segmentation in multiple sclerosis: a systematic review and meta-analysis. Lancet Neurol. 2017;16(10):797–812. doi:10.1016/S1474-4422(17)30278-8

34. Kupersmith MJ, Garvin MK, Wang JK, Durbin M, Kardon R. Retinal ganglion cell layer thinning within one month of presentation for optic neuritis. Mult Scler. 2016;22(5):641–648. doi:10.1177/1352458515598020

35. Saidha S, Syc SB, Ibrahim MA, et al. Primary retinal pathology in multiple sclerosis as detected by optical coherence tomography [published correction appears in Brain. 2013 Dec;136(Pt 12):e263]. Brain. 2011;134(Pt 2):518–533. doi:10.1093/brain/awq346

36. Huang-Link YM, Al-Hawasi A, Lindehammar H. Acute optic neuritis: retinal ganglion cell loss precedes retinal nerve fiber thinning. Neurol Sci. 2015;36(4):617–620. doi:10.1007/s10072-014-1982-3

37. Tienari PJ, Salonen O, Wikström J, Valanne L, Palo J. Familial multiple sclerosis: MRI findings in clinically affected and unaffected siblings. J Neurol Neurosurg Psychiatry. 1992;55(10):883–886. doi:10.1136/jnnp.55.10.883

38. Siger-Zajdel M, Filippi M, Selmaj K. MTR discloses subtle changes in the normal-appearing tissue from relatives of patients with MS. Neurology. 2002;58(2):317–320. doi:10.1212/wnl.58.2.317

39. De Stefano N, Cocco E, Lai M, et al. Imaging brain damage in first-degree relatives of sporadic and familial multiple sclerosis. Ann Neurol. 2006;59(4):634–639. doi:10.1002/ana.20767

40. Ehtesham N, Rafie MZ, Mosallaei M. The global prevalence of familial multiple sclerosis: an updated systematic review and meta-analysis. BMC Neurol. 2021;21(1):246. Published 2021 Jun 28. doi:10.1186/s12883-021-02267-9

